# Assessing the contribution of gut-to-lung translocation to bacterial colonization and antibiotic resistance in an ICU patient

**DOI:** 10.1101/2022.01.17.22269403

**Authors:** Rachel M Wheatley, Julio Diaz Caballero, Thomas E. van der Schalk, Fien HR De Winter, Natalia Kapel, Claudia Recanatini, Leen Timbermont, Jan Kluytmans, Mark Esser, Alicia Lacoma, Cristina Prat-Aymerich, Antonio Oliver, Samir Kumar-Singh, Surbhi Malhotra-Kumar, R. Craig MacLean, WP3A working group

**Author notes:** These authors contributed equally (Rachel M Wheatley, Julio Diaz Caballero).

## Abstract

Bacteria have the potential to migrate between sites in the human body, but the dynamics and consequences of within-host translocation remain poorly understood. Here we investigate the link between gut and lung *Pseudomonas aeruginosa* populations in an intensively sampled ICU patient using a combination of genomics, isolate phenotyping, host immunity profiling, and clinical data. Crucially, we show that lung colonization was driven by the repeated translocation of bacterial clones from the gut. Meropenem treatment for a suspected urinary tract infection selected for elevated resistance in both the gut and lung. However, resistance was driven by parallel evolution and organ-specific selective pressures, and within-host transmission had only a minor impact on AMR. These findings suggest that reducing intestinal colonization of *Pseudomonas* may be an effective way to prevent lung infections in critically ill patients.

## Introduction

Bacteria often colonize multiple anatomical sites in human hosts, but the dynamics of within-host translocation and its consequences for pathogenesis and host adaptation remain poorly understood^1-3^. For example, advances in microbiome profiling methods have shown that the gut microbiome can transmit to the lungs of critically ill patients^4,5^, and translocation is associated with poorer outcomes in mechanically ventilated patients^6^. While gut-to-lung translocation has been demonstrated at the microbiome level, the dynamics and consequences of translocation for individual pathogens remain poorly understood.

*P. aeruginosa* is an opportunistic pathogen that is a major cause of healthcare-associated infections worldwide^7,8^, most notably in patients with compromised immunity^9,10^. *Pseudomonas* is not considered to be a typical member of the gut microbiome, and intestinal colonization with *Pseudomonas* is associated with an increased risk of developing lung infections^11-13^ and mortality^14^. Gut colonization usually precedes lung infection, and the same strain is often found in the gut and lungs, suggesting that the gut acts as a reservoir of *Pseudomonas* that can be transmitted to the lung and other infection sites^15-17^. However, direct evidence for gut-to-lung transmission of *P. aeruginosa* is lacking, and it is possible that intestinal carriage simply reflects an innate susceptibility to *Pseudomonas* infection or proximity to a source of *Pseudomonas* that can independently colonize the lung and gut.

To test the importance of gut-to-lung transmission in *Pseudomonas* colonization and antimicrobial resistance (AMR), we carried out an in-depth case study on a single intensively sampled ICU patient over a 30-day period. We used phylogenetic approaches to test for transmission, and a combination of genomic and phenotypic methods to study the link between AMR and within-host transmission.

## Results

### Clinical timeline

The focal patient was admitted to ICU of Hospital Universitari Germans Trias i Pujol in Badalona, Spain with a primary diagnosis of seizure. The patient was immediately treated with amoxicillin clavulanate, which is not active against *P. aeruginosa*, due to suspected aspiration of oropharyngeal or gastric contents into the lower respiratory tract. The patient was enrolled in ASPIRE-ICU trial^18^ at 48hrs post admission (hereafter day 1) and lung *P. aeruginosa* colonization was detected. Gut colonization was detected following meropenem treatment for a suspected urinary tract infection at day 12, and meropenem resistant *P. aeruginosa* ultimately colonized the lung. This complex clinical timeline suggests that translocation between the gut and lung may have occurred (Figure 1), but clinical data and isolate phenotypes alone provide limited insights into the underlying drivers of within-host transmission and AMR.

**Figure 1:**
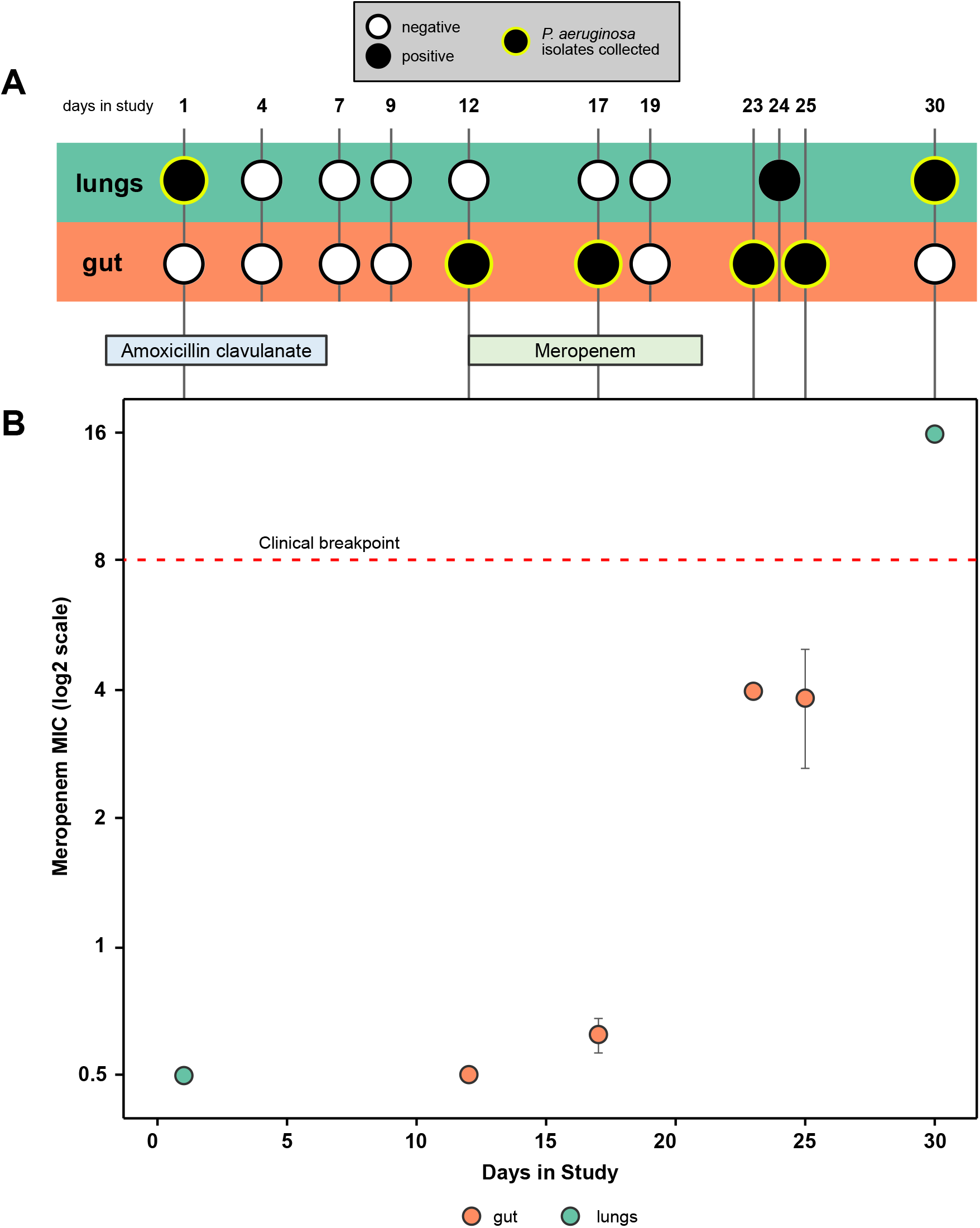
Clinical timeline and resistance phenotyping. (A) Timeline of patient sampling, showing samples that tested positive or negative for *P. aeruginosa* colonization. Sampling points from which isolates were collected are highlighted with a green ring. The patient was treated with Amoxicillin clavulanate from 2 days prior to enrolment until day 6 and with Meropenem from day 12 to day 21. (B) Meropenem minimum inhibitory concentration (MIC) (mean +/- standard error) for isolates from the gut (green) and lung (purple). Meropenem resistance increased over time, and *P. aeruginosa* isolates from the final lung sample were above the EUCAST clinical breakpoint for meropenem resistance (red dashed line). Amoxicillin clavulanate resistance was not measured as this antibiotic is not active against *P. aeruginosa*.

### Genomic insights into pathogen colonization and evolution

To characterize the genetic diversity within this patient, we used long and short read sequencing to construct a hybrid assembly for a single isolate, yielding a ∼6.3Mb ST782 reference genome distributed across 5 contigs. Short-read sequences of lung (n=12) and gut (n=40) isolates were mapped to this reference genome, and we identified polymorphic SNPs (n=17), indels (n=7), and variation in presence/absence of a 190kb genomic island (Supplementary Table 1; Supplementary Figure 1).

The genetic diversity found in this patient could reflect either (i) recurrent colonization/infection by multiple clones or (ii) within-host evolution of a single clone. To discriminate between these processes, we reconstructed the phylogeny isolates using *P. aeruginosa* PA1, a closely related ST782 genome, as an outgroup (Figure 2A-D). The number of variants per isolate correlated strongly with the day of isolate collection (Figure 2E; r^2^=.62, F_1,50_=82, P<.0001), supporting the idea that the within-patient diversity was driven by the evolution of a single cell that colonized the patient approximately 2 weeks prior to ICU admission while the patient was in the community (predicted MRCA at day -15 +/- 3 days). No other patients within the ASPIRE-ICU cohort at this hospital were colonized by *P. aeruginosa* ST782 during the trial, providing further support for the within-host diversification as opposed to repeated colonization. The rate of evolution in this patient was 51 SNPs/year (standard error (s.e.)=5.84, t=9.07, P<.0001), which is higher than the typical evolutionary rate of bacterial pathogens of 1-10 SNPs/year^3^. However, this elevated evolutionary rate is only ∼2 fold greater than the rate reported from another patient in this trial^19^, highlighting the high *in vivo* mutation rate of *P. aeruginosa* in critically ill patients.

**Figure 2:**
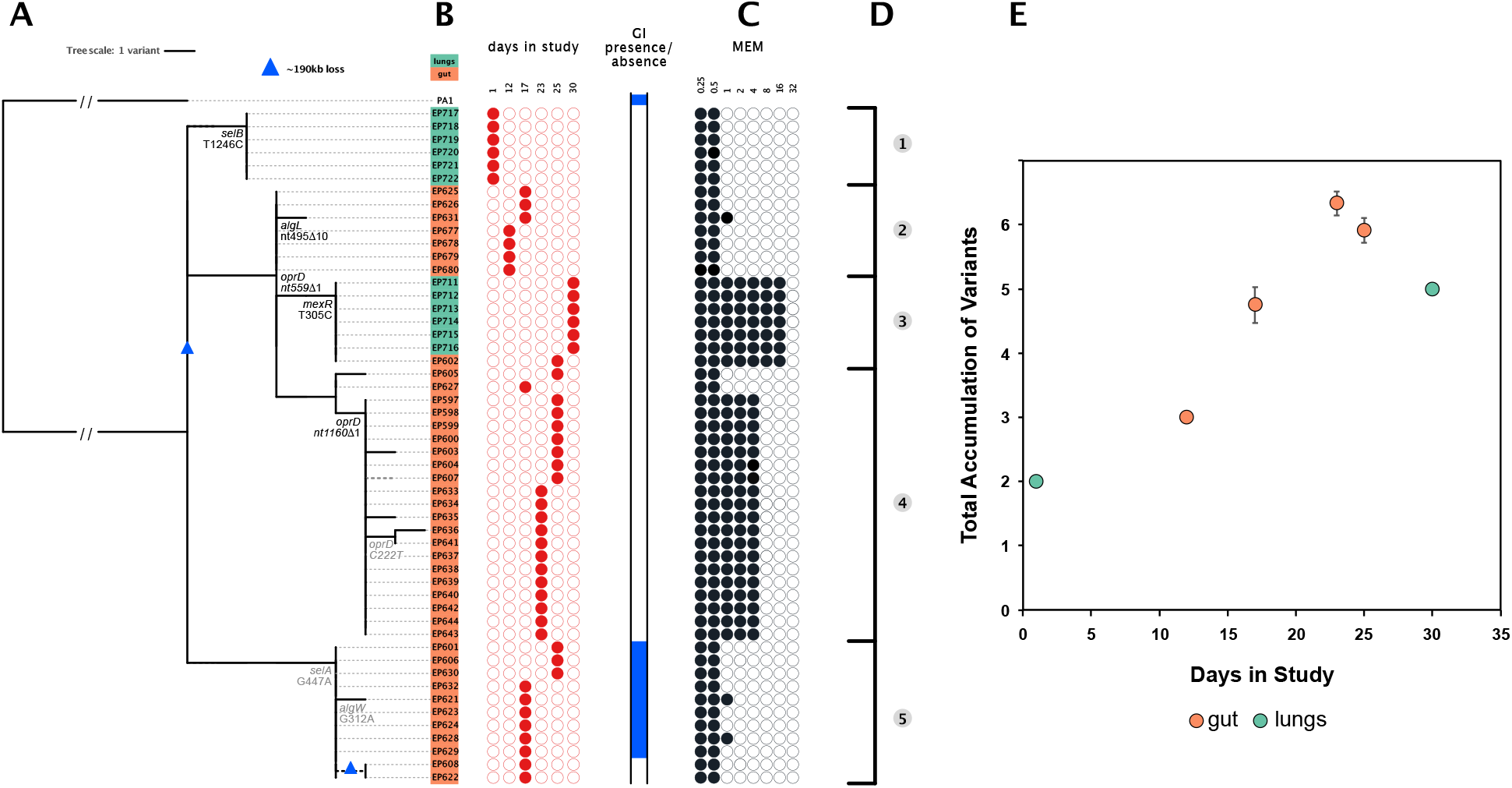
Genome sequencing and phylogenetic analysis. (A) Phylogenetic reconstruction of lung (n=12) and gut (n=40) isolates rooted using *P. aeruginosa* PA1, another ST782 isolate, as the outgroup. Putatively adaptive polymorphisms in genes or pathways showing parallel evolution are annotated on the phylogeny and silent mutations are shown in light grey. A polymorphism in a known multi-drug efflux pump regulator (*mexR*) is also highlighted.

Variation in the presence/absence of a 190kB genomic island is shown, and inferred losses of the genomic island are identified with blue triangles in the tree. (B) Isolate name, lung (green) or gut (orange) origin, and day in study of collection.n (C) Susceptibility to meropenem (MEM) for each isolate is presented with filled black circles in log_2_ scale of the minimum inhibitory concentration (MIC). (D) The topology of the tree suggested five distinct groups based on the identification of small polymorphisms. (E) Rate of accumulation of variants (mean +/- s.e) as a function of time per body site suggests within-host evolution of a clone rather than recurrent episodes of colonization.

Signatures of parallel evolution provide a simple way to identify putative beneficial mutations that underpin adaptation to the novel environment of the human host^3,20^. Parallel evolution occurred in 3 genes or operons that have functional roles in resistance to carbapenem antibiotics (*oprD*)^20^, alginate biosynthesis (*algW, algL*)^21^, and selenocysteine biosynthesis (*selA and selB*)^22^. These putative pathoadaptive mutations accounted for 7 of the 24 variants, providing strong evidence for rapid adaptation to the host environment. Interestingly, 3 of these 7 mutations were synonymous, suggesting that transcription efficiency may have been a key target of selection^23^. The variable genomic island was lost on two independent occasions, suggesting that loss of this element was adaptive. Inferring the selective advantage of large scale deletions is difficult, but it is worth noting that this island carries pyoverdine biosynthesis genes that are selected against in the host environment^24^.

Bacterial phylogenies are a powerful tool to detect transmission events^3,25,26^. In this case, the most parsimonious explanation for the distribution of lung and gut isolates on the phylogeny is that *Pseudomonas* colonized the gut of the patient and then transmitted to the lung on at least two independent occasions (Figure 2). Initial lung colonization was driven by the gut to lung transmission of a lineage that acquired mutations in *gslA* and *selB* (Figure 2A – lineage 1). The lack of diversity within this lineage suggests that this transmission event was driven by the outgrowth of a single cell soon before the patient was enrolled in the trial, possibly as a result of the broncoaspiration upon admission to ICU (i.e., at day -1). Secondary lung colonization (Figure 2A – lineage 3) was caused by the growth of a clone with mutations in the *oprD* porin, which is a key carbapenem sensitivity determinant, and *mexR*, which regulates the expression the MexAB-OprM multi-drug efflux pump^27^ (Figure 2A – lineage 2). This *oprD/mexR*T305C lineage is nested within a broader clade of gut isolates, providing strong evidence that secondary lung colonization was driven by a second gut to lung transmission event likely occurred at some point after day 12.

### Immune response to lung colonization

Lung colonization provides *P. aeruginosa* with the opportunity to establish infection by adhering to the mucosal surface and penetrating the epithelial barrier, leading to the development of pneumonia. To investigate the role of host immunity in preventing infection, we measured the abundance of a panel of host immune effectors in samples of endotracheal aspirate (ETA) (Figure 3). Secondary lung colonization (day 17-19 samples) was associated with a host immune response that is indicative of a homeostatic and healing state, with high levels of expression of IL-33, Fractalkine, and IL-4, which have previously been to shown to enhance the clearance of *P. aeruginosa*^28^. Colonization was also associated with a >10-fold increase in the concentration of IL-22, which protects against infections caused by attaching and effacing bacterial pathogens by increasing mucous production and by limiting excessive inflammation mediated by neutrophil influx^29,30^. Measuring cytokine levels in ETA samples from the lungs provides a direct measurement of immune response, but one concern over this approach is that it is possible for individual samples to give high concentrations of all cytokines, for example as a result of patient dehydration. However, in this case levels of IL-8 remained essentially constant across samples, supporting the idea spikes of protective cytokines were not an artefact.

**Figure 3:**
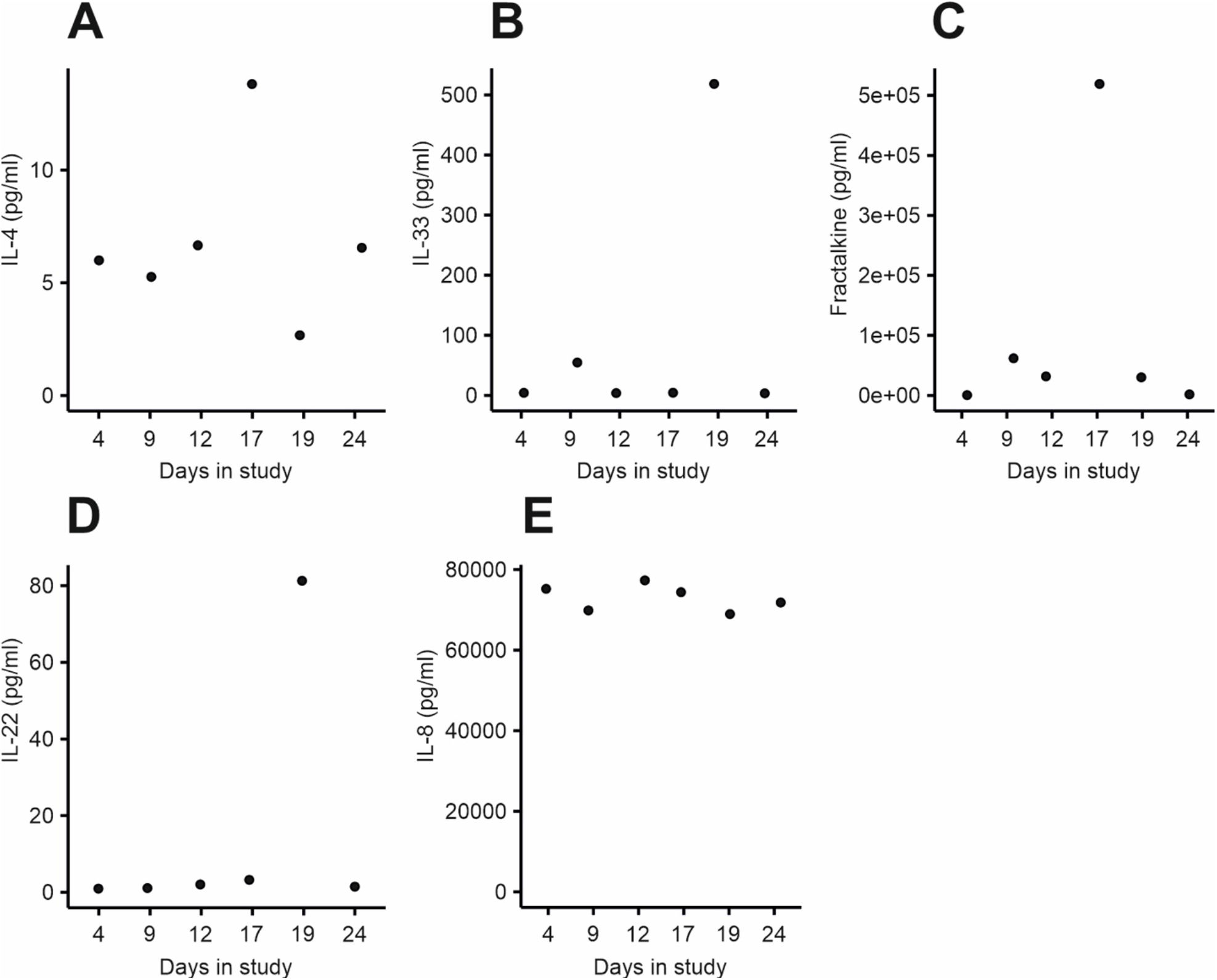
Cytokine concentrations were measured in ETA samples collected over the course of the study at days 4, 9, 12, 17, 19 and 24. The following cytokines were measured: (A) IL-4, (B) IL-33, (C), Fractalkine, (D) IL-22, (E) IL-8.

### Drivers of antibiotic resistance

*P. aeruginosa* has high levels of intrinsic antibiotic resistance and a remarkable ability to evolve increased resistance under antibiotic treatment^31,32^. Given the possibility of migration between the gut and lung, we next sought to understand the relative contributions of migration, mutation and selection to the origin and spread of meropenem resistance in this patient. The phylogeny clearly shows that elevated meropenem resistance evolved on 2 separate occasions due to mutations in *oprD* and *mexR* (Figure 2, lineages 4 and 5). However, it is challenging to follow the dynamics of meropenem resistance mutations using isolates alone due to the limited number of isolates sequenced (n=52) and the gaps in the sampling of isolates. To overcome this problem, we combined isolate sequencing data with amplicon sequencing of *oprD* using DNA extracted directly from ETA samples, some of which were not screened for isolates according to ASPIRE-ICU protocol (Figure 4A-B).

**Figure 4:**
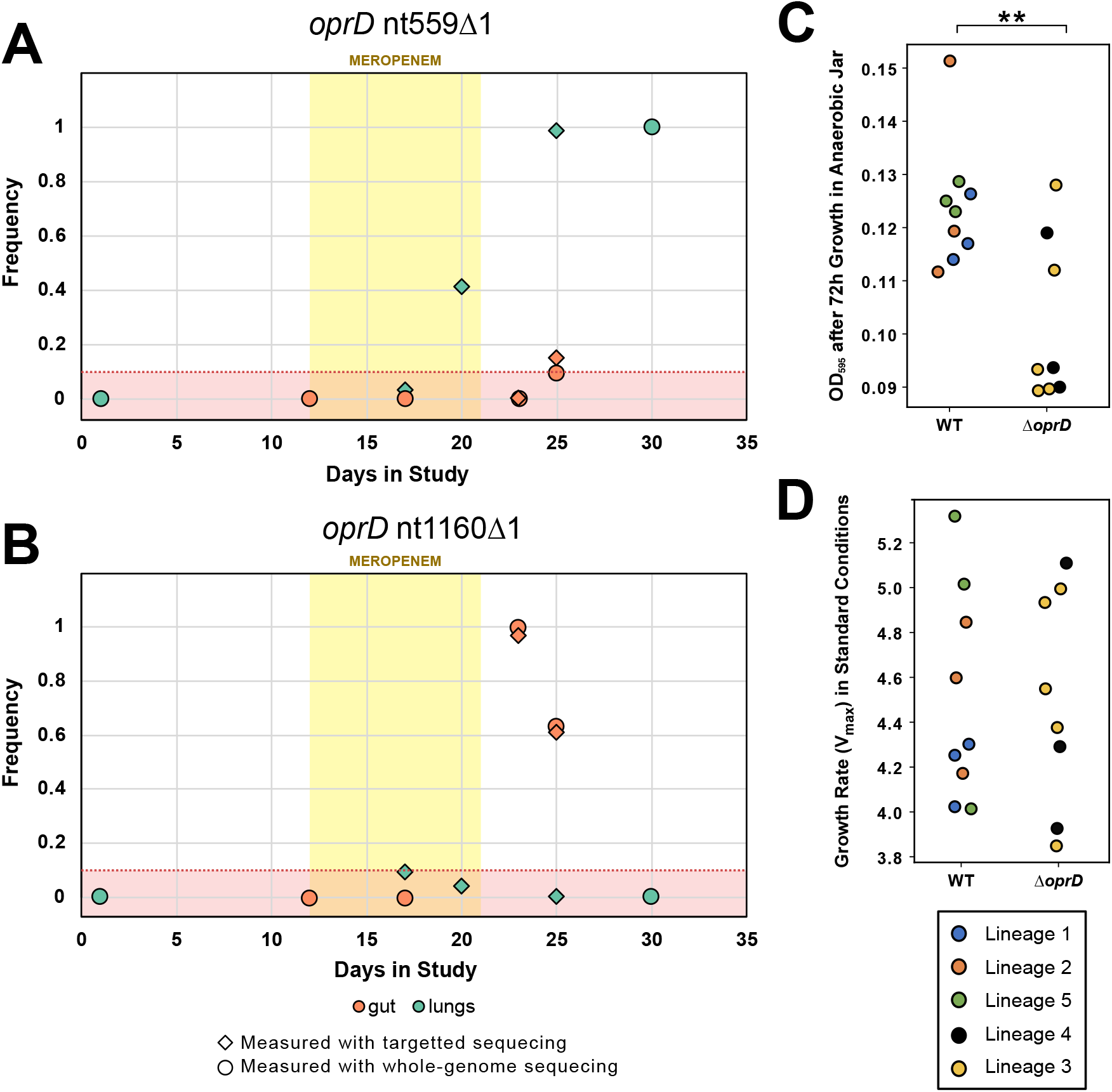
Evolution and transmission of meropenem resistance. (A and B) Dynamics of *oprD* variants, as determined by combining isolate WGS (circle) and *oprD* amplicon (diamond) sequencing data. The yellow area shows the window of meropenem treatment and the red area shows the minimum detection limit of variants from amplicon sequencing due to the high error rate of nanopore sequencing. The *oprD* nt559Δ1 variant was detected in a single gut isolate at day 25 (frequency=1/12) and the frequency of this variant in amplicons was marginally greater than the detection threshold (frequency = 97/671). (C and D) Growth of isolates with an o*prD* variant (Δ o*prD*) compared to isolates with the wild-type *oprD* background. Anaerobic growth (C) was measured as OD595 after 72 hours growth in anaerobic broth. Aerobic growth (D) was measured as exponential growth rate in standard culture conditions. Isolates are colour coded according to phylogenetic lineage, as defined in Figure 2. *oprD* mutations were associated with impaired growth under anaerobic conditions (P=.010), but not aerobic conditions (P=.950), as judged by a nested ANOVA. Data from isolates from different lineages are shown together because fitness measures did not differ between lineages nested within *oprD* genotype (P >.5).

The most common *oprD* mutation in sequenced isolates (nt1160Δ1) arose in the main clade of intestinal isolates (Figure 2, lineage 4) and it was never convincingly detected in lung samples (Figure 4B), providing good evidence that this mutation arose in the gut and swept to high frequency under meropenem treatment. Isolate sequencing revealed a second frameshift mutation in *oprD* (nt559Δ1) that was linked to a mutation in the *mexR* effux pump transcriptional regulator. Isolate sequencing revealed the presence of the *oprD* nt559Δ1/*mexR*T305C lineage in gut samples from day 25 and lung samples from day 30, suggesting that that this lineage may have evolved resistance to meropenem in the gut and then transmitted to the lung. However, amplicon sequencing revealed that *oprD* nt559Δ1 swept to fixation in the lung during meropenem treatment (Figure 4A). This mutation was detected in gut isolates and amplicons that were sampled after meropenem treatment had ended (i.e. day 25), providing good evidence that the *oprD* nt559Δ1 mutation arose in the lung, and secondarily transmitted to the gut (Figure 4A).

Antibiotic concentrations vary between host tissues, and it is unclear to what extent pathogen populations adapt to local variation in selective pressures associated with antibiotic treatment. Meropenem achieves higher concentrations in lung tissues than in the gut^33^, suggesting that selection for resistance should be stronger in the lung than in the gut. Consistent with this expectation, isolates from the lung-associated *oprD* nt559Δ1/*mexR*T305C lineage had higher meropenem resistance (MIC=16 mg/L, s.e.=0 mg/L, n=19) than isolates from the gut-associated *oprD* nt1160Δ1 lineage (MIC=4 mg/L, s.e.=0 mg/L, n=7; Figure 2C), demonstrating organ-specific adaptation to underlying differences in meropenem concentration.

A key challenge in evolutionary studies of AMR is to understand how resistance can be maintained in pathogen populations in the absence of continued antibiotic use^34,35^. In this case, *oprD* nt559Δ1 resistance remained stable in the lung following antibiotic treatment, but the frequency of *oprD* nt1160Δ1 declined in the gut due to the expansion of a carbapenem sensitive lineage (Figure 2A – lineage 5). We speculate that this lineage may have survived carbapenem treatment by either colonizing a region of the gut with low carbapenem toxicity or by forming persister cells.

To test the role of selection in the stability of resistance, we measured the growth rate of isolates of all 5 major phylogenetic lineages in anaerobic culture medium (Figure 4C) and aerobic culture medium (Figure 4D), which recapitulates one of the physiological differences between the gut and lung. Meropenem resistant lineages (Δ o*prD*) were not associated with decreased growth rate under aerobic conditions, suggesting that the *oprD* and *mexR* mutations have little, if any, associated costs under these conditions (Figure 4D; F_1,12_=.0032, P=.956). In contrast, both meropenem lineages were associated with decreased growth under anaerobic conditions, suggesting that fitness costs associated with *oprD* mutations drove the loss of resistance in the gut (Figure 4C; F_1,12_=9.27, P=.010).

## Conclusion

The goal of this project was to understand the link between gut and lung *Pseudomonas* colonization in a single patient. By combining clinical and genomic data, we were able to demonstrate that transmission from the gut caused repeated lung colonization. Whilst it is difficult to generalize the findings of a single case study, these findings support the idea that gut to lung transmission is a major driver of *P. aeruginosa* respiratory tract colonization in critically ill patients^11,12,36^. Lung colonization was associated with the production of cytokines that protect against *P. aeruginosa* infection, suggesting that a robust host immune response is key to preventing *P. aeruginosa* lung infection following lung colonization.

Carbapenem antibiotics such as meropenem are key to the treatment of *P. aeruginosa* infections^19,37,38^, and carbapenem-resistant *P. aeruginosa* has been identified as an important threat by the World Health Organisation and the Centers for Disease Control and Prevention. In this patient, meropenem treatment for a suspected urinary tract infection drove the repeated evolution of resistance, providing a poignant example of the importance of ‘bystander selection’ for AMR^39^. Ultimately, selection led to the emergence of a stable population of highly resistant bacteria in the lung, suggesting that the respiratory tract can act as a source of carbapenem resistant *Pseudomonas* that can transmit to other body sites and potentially to other patients.

Migration increases genetic variation^40^, suggesting that within-host transmission may accelerate bacterial adaptation to antibiotics^1-3,41-44^. In this case resistance was driven by local mutation and selection within organs, leading to the emergence of a highly structured pathogen population^1-3,43^ shaped by a mosaic of selective pressures stemming from antibiotic treatment. We speculate that the high *in vivo* mutation rate of *Pseudomonas* was key to shaping local adaptation to antibiotic selection across tissues, and that within-host transmission is likely to provide a more important source of resistance at smaller spatial scales^2^, or when mutation rate is low.

Hospital acquired infections caused by epidemically successful MDR and XDR strains of *P. aeruginosa* have become a serious problem worldwide^45^, and there is an urgent need to develop new antibiotics to treat infections caused by these strains. At the same time, the incredible ability of *Pseudomonas* to evolve resistance to antibiotic treatment^19,31,32,41^ highlights the need to develop novel approaches to prevent or treat *Pseudomonas* infections. Our study suggests that preventing gut colonization or gut to lung transmission may be an effective strategy for preventing *Pseudomonas* infection in critically ill patients^46-49^.

## Materials and methods

### Clinical timeline

The patient was admitted to the intensive care unit (ICU) of Hospital Universitari Germans Trias i Pujol in Badalona, Spain with a primary diagnosis of seizure. At the time of ICU admission, the patient did not suffer from pneumonia or any other active *P. aeruginosa* infection (APACHE-II score = 22 and Glasgow Coma scale = 3). No antibiotic use was reported in the two weeks prior to hospital admission. After 48 hours of ICU admission, informed consent was obtained and the patient was enrolled in the ASPIRE-ICU study (day 1)^18^. Mechanical ventilation (MV) was started on ICU admission and was continued for a total duration of 39 days. Amoxicillin/clavulanic acid (1000 mg IV q8h for 8 days) was started on ICU admission for bronchoaspiration, the suspected inhalation of oropharyngeal or gastric contents into the lower respiratory tract. Meropenem (1000 mg IV q8h for 10 days) was started on day 12 to treat a suspected urinary tract infection. Patient samples were collected and screened for *P. aeruginosa* isolates as described in^19^ until day 30, and the patient was discharged from the ICU and transferred to a general medical ward on day 41.

### Resistance phenotyping

All isolates were grown from glycerol stocks on Luria-Bertani (LB) Miller Agar plates overnight at 37 °C. Single colonies were then inoculated into LB Miller broth for 18–20 h overnight growth at 37 °C with shaking at 225 rpm. Overnight suspensions were serial diluted to ∼5 × 10^5^ CFU/mL. Resistance phenotyping to meropenem was carried out as minimum inhibitory concentration (MIC) testing via broth microdilution as defined by EUCAST recommendations^50,51^, with the alteration of LB Miller broth for growth media and the use of *P. aeruginosa* PAO1 as a reference strain. Resistance to meropenem was assayed along a 2-fold dilution series between 0.25 ug/mL - 64 ug/mL. We defined growth inhibition as OD_595_ < 0.200 and we calculated the MIC of each isolate as the median MIC score from three biologically independent assays of each isolate (Supplementary Table 2).

### Growth assays

*P. aeruginosa* isolates were grown from glycerol stocks on LB Miller Agar plates overnight at 37 °C. Single colonies were then inoculated into LB Miller broth for 18–20 h overnight growth at 37 °C with shaking at 225 rpm. Overnight suspensions were serially diluted to an OD_595_ of ∼0.05 within the inner 60 wells of a 96-well plate equipped with a lid. To assess growth rate under standard aerobic conditions, isolates were then grown in LB Miller broth at 37 °C and optical density (OD595nm) measurements were taken at 10-min intervals in a BioTek Synergy 2 microplate reader set to moderate continuous shaking. Growth rate (Vmax; mOD/min) was calculated as the maximum slope of OD versus time over an interval of ten consecutive readings, and we visually inspected plots to confirm that this captured log-phase growth rate. We measured the growth rate of all 52 gut and lung isolates with a minimum of seven biological replicates to assess the relationship between meropenem resistance and fitness (Supplementary Table 2). We measured anaerobic growth using an anaerobic jar (Thermo Scientific^™^ Oxoid^™^ AnaeroJar^™^ base jar) system with anaerobic gas generating sachets (Thermo Scientific^™^ Oxoid^™^ AnaeroGen^™^ sachets). An Oxoid Resazurin indicator strip was placed in the jar as an indicator to confirm generation of an anaerobic environment. For growth measurements, single colonies were inoculated into LB Miller broth in the wells of a 96-well plate and placed in the anaerobic jar for 72 hours, after which plates were removed and OD_595_ was measured. For the comparison of o*prD* variant (Δ o*prD*) isolates to wild-type *oprD* background (WT *oprD*) isolates, growth measurements were taken for a minimum of three isolates (and a minimum of three biological replicates) selected as representatives from each phylogeny group to generate a mean growth measurement for each Δ o*prD* and WT *oprD* group. To test for an association between *oprD* mutations and impaired growth we used a nested ANOVA that included main effects of *oprD* (ie either WT or Δ o*prD*, 1 df) and phylogenetic lineage nested within *oprD* (5 lineages shown in Figure 2, 3 df).

### Illumina sequencing

All isolates were sequenced in the MiSeq or NextSeq illumina platforms yielding a sequencing coverage of 21X–142X. Raw reads were quality controlled with the ILLUMINACLIP (2:30:10) and SLIDINGWINDOW (4:15) in trimmomatic v. 0.39^52^. Quality controlled reads were assembled for each isolate with SPAdes v. 3.13.1^53^ with default parameters. These assemblies were further polished using pilon v. 1.23^54^ with minimum number of flank bases of 10, gap margin of 100,000, and kmer size of 47. Resulting contigs were annotated based on the P. aeruginosa strain UCBPP-PA14^55^ in prokka v. 1.14.0^56^. Each isolate was typed using the Pseudomonas aeruginosa multi-locus sequence typing (MLST) scheme from PubMLST (last accessed on 11.06.2021)^57^.

### Long-read sequencing

Two isolates (EP717 and EP623) were sequenced using the Oxford nanopore MinION platform with a FLO-MIN106 flow-cell and SQK-LSK109 sequencing kit. EP717 had sequencing coverage of 141X and EP623 of 233X. Raw reads were basecalled using guppy v. 0.0.0+7969d57 and reads were demultiplexed using qcat v. 1.1.0 (https://github.com/nanoporetech/qcat). Resulting sequencing reads were assembled using unicycler v. 0.4.8^58^, which used SAMtools v. 1.9^59^, pilon v. 1.23^54^, and bowtie2 v. 2.3.5.1^60^, in hybrid mode with respective illumina reads. The EP717 assembly had a N50 of 1,797,327 for a total of 6,217,789 bases distributed in 11 contigs. The EP623 assembly had a N50 of 6,133,283 for a total of 6,330,243 bases distributed in 5 contigs.

### Variant calling

To identify mutations and gene gain/loss during the infection, short-length sequencing reads from each isolate were mapped to each of the long-read de novo assemblies with BWA v. 0.7.17 ^61^ using the BWA-MEM algorithm. Preliminary SNPs were identified with SAMtools and BCFtools v. 1.9. Low-quality SNPs were filtered out using a two-step SNP calling pipeline, which first identified potential SNPs using the following criteria: 1) Variant Phred quality score of 30 or higher, 2) At least 150 bases away from contig edge or indel, and 3) 20 or more sequencing reads covering the potential SNP position. In the second step, each preliminary SNP was reviewed for evidence of support for the reference or the variant base; at least 80% of reads of Phred quality score of 25 or higher were required to support the final call. An ambiguous call was defined as one with not enough support for the reference or the variant, and, in total, only one non-phylogenetically informative SNP position had ambiguous calls. Indels were identified by the overlap between the HaplotypeCaller of GATK v. 4.1.3.0^62^ and breseq v. 0.34.0^63^. The variable genome was surveyed using GenAPI v. 1.098^64^ based on the prokka annotation of the short-read de novo assemblies. The presence or absence of genes in the potential variable genome was reviewed by mapping the sequencing reads to the respective genes with BWA v.0.7.17^61^.

Patient sample DNA was derived from the ETA samples. Similar to the culture of these samples, samples were blended (30,000 rpm, probe size 8 mm, steps of 10 s, max 60 s in total), diluted 1:1 v/v with Lysomucil (10% Acetylcysteine solution) (Zambon SA, Belgium) and incubated for 30 min at 37 °C with 10 s vortexing every 15 min^19^. Thereafter, 250µl of the liquefied sample was used in the ZymoBIOMICS DNA Miniprep Kit (Zymo Research, CA USA) for DNA extraction.

### Amplicon Sequencing of oprD

Amplicon sequencing of the *oprD* gene was carried out to quantify the presence of the two key *oprD* variants observed in the isolate sequencing in whole gDNA samples that were available from the lung and gut of this patient. Patient samples were blended (30,000 rpm, probe size 8 mm, steps of 10 s, max 60 s in total), diluted 1:1 v/v with Lysomucil (10% Acetylcysteine solution) (Zambon SA, Belgium) and incubated for 30 min at 37 °C with 10 s vortexing every 15 min, as described in^19^. Thereafter, 250µl of the liquefied sample was utilised in the ZymoBIOMICS DNA Miniprep Kit (Zymo Research, CA USA) for DNA extraction.

A PCR amplification strategy using barcoded primers to amplify the *oprD* gene (1489bp product length) and add sample specific DNA barcodes was followed as described in^65^. The method used a universal reverse primer and sample specific forwards primers containing 12nt barcodes (Supplementary Table 3)^65^. The barcoded *oprD* PCR products were pooled and sequenced on an Oxford nanopore MinION platform using a FLO-MIN106 flow-cell and the SQK-LSK109 Ligation Sequencing kit. Amplicon sequencing raw reads were basecalled using guppy v. 0.0.0+7969d57. This yielded 163,766 reads with an estimated read length N50 of 2.64kb. The data was demultiplexed allowing 2/12 sequencing errors in the barcode sequence and a maximum of 1 error in the downstream and upstream 4-mer. To identify the genotype of each read, we searched for the 11-mer sequence including the variant base and 5 bases downstream and upstream from this position. Using this conservative approach, we recovered 32 - 43% of the reads (Supplementary Table 4).

### Cytokine measurements

Levels of interleukin (IL-)4, IL-33, IL-22, IL-8 and fractalkine were measured in ETA as previously described^19^. Briefly, ETA was measured in a final dilution of 1/16 on a U-plex panel (Mesoscale Discovery, Rockville, MD, USA) following the manufacturer’s instructions. The plate was coated with capturing antibodies for 1 h with shaking incubation at room temperature followed by washing off the plate. Samples were loaded and incubated for 1 h, after which the plate was washed and incubated with detection antibodies for 1 h. A final wash was performed and MSD gold reading buffer was applied before reading the plate in the QuickPlex^™^ SQ 120 (MSD).

## Data Availability

All data produced in the present study are available upon reasonable request to the authors.

## Supplementary Information

**Supplementary Figure 1:** Identification of genomic island. The mapping of Illumina and Oxford Nanopore sequencing data to the reference P. aeruginosa PA1 revealed isolates (e.g. EP608) that lacked a genomic region of ∼125 kb. In addition, the depth of coverage of the ∼75 kb upstream region was 2X greater in isolates carrying the ∼125 kb region (e.g. EP623). This information revealed a ∼200 kb genomic region, which carried a second copy of the ∼75 kb region and additional ∼125 kb genetic content.

**Supplementary Table 1:** Genomic variants

**Supplementary Table 2:** Metadata for the 52 P. *aeruginosa* isolates collected in study, including meropenem MICs and growth measurements

**Supplementary Table 3:** Primer table

**Supplementary Table 4:** Reads recovered from amplicon sequencing experiments

## Acknowledgements

This research was supported by Wellcome Trust Grant (106918/Z/15/Z) and the Innovative Medicines Initiative Joint Undertaking under COMBACTE-MAGNET (Combatting Bacterial Resistance in Europe-Molecules against Gram-negative Infections, grant agreement no. 115737) and COMBACTE-NET (Combatting Bacterial Resistance in Europe-Networks, grant agreement no. 115523), resources of which are composed of financial contribution from the European Union’s Seventh Framework Program (FP7/2007-2013) and EFPIA companies’ in kind contribution. We thank the Oxford Genomics Center (funded by Wellcome Trust Grant 203141/Z/16/Z) for the generation and initial processing of Illumina sequence data. We thank the local ASPIRE ICU research team for their contribution to this project at Hospital Universitari Germans Trias i Pujol.

## Ethics statement

The study protocol was approved by the Research Ethics Committee of the Germans Trias i Pujol University Hospital. This study was conducted according to the principles of the Declaration of Helsinki, in accordance with the Medical Research Involving Human Subjects Act and local guidelines in the participating countries.

